# Non-Consensual Sex among Japanese Women in the COVID-19 Pandemic: A Large-Scale Nationwide Survey-Based Study

**DOI:** 10.1101/2024.02.16.24302967

**Authors:** Tomoya Suzuki, Yudai Kaneda, Yasuhiro Kotera, Akihiko Ozaki, Tetsuya Tanimoto, Divya Bhandari, Sayaka Horiuchi, Takahiro Tabuchi

## Abstract

**Background:** Non-consensual sex including rape and sexual assault has been a global concern and may have been influenced by the COVID-19 pandemic, however the information on this topic is limited. Therefore, our objective was to survey the incidence rate of non-consensual sex among Japanese women aged 15-79 years between April to September 2020, following the COVID-19 pandemic in Japan.

**Materials and Methods:** We utilized the data obtained from a nationwide, cross-sectional internet survey conducted in Japan between August and September 2020. Sampling weights were applied to calculate national estimates, and multivariable logistic regression was performed to identify factors associated with non-consensual sex. Data was extracted from a cross-sectional, web-based, self-administered survey of approximately 2.2 million individuals from the general public, including in men and women.

**Results:** Excluding men and responses with inconsistencies, the final analysis included 12,809 women participants, with 138 (1.1%) reporting experiencing non-consensual sex within a five-month period. Being aged 15–29 years and having a worsened mental or economic status were associated with experiencing non-consensual sex.

**Conclusions:** Early intervention to prevent individuals from becoming victims of sexual harm should be extended to economically vulnerable and young women, especially during times of societal upheaval such as the COVID-19 pandemic. Additionally, Japan should prioritize the implementation of comprehensive education on the concept of sexual consent.

## INTRODUCTION

Non-consensual sex refers to engaging in sexual behavior without obtaining consent to touch, observe, or perform sexual acts involving private body parts that exceed the boundaries of another person’s body or mind (1,2). Non-consensual sex can take many forms, including rape, transactional sex, cross-generational sex, unwanted touch, and molestation. In the European Union, between 45% to 55% of women have experienced sexual harassment since the age of 15 (3). Previous research has identified common factors associated with victims, including ages between 10 and 30 years, mental vulnerability, and financial hardship (4). Women with these characteristics are at a higher risk of experiencing non-consensual sex compared to those without these traits (4). Furthermore, women with these characteristics often find it challenging to disclose incidents of non-consensual sex to others, making it difficult to address these issues (5).

Of note, non-consensual sex is reported to increase due to disruptions in social conditions, such as natural disasters and worsened economic conditions. For instance, incidents of non-consensual sex against women dramatically increased during times of economic and psychological turmoil, such as the 2005 Hurricane Katrina in the United States and the 2011 Great East Japan Earthquake (6–8). Therefore, it is crucial to investigate the prevalence of non-consensual sex after the COVID-19 pandemic.

At the onset of the COVID-19 pandemic, it was reported that 243 million girls and women aged 15-49 years had been subjected to non-consensual sex worldwide (9). This was attributed to the increase in the mandatory stay-at-home measure due to lockdowns, and resulting in reduced access to social support from teachers, friends and caregivers (10). Awareness of the status of non-consensual sex during the COVID-19 pandemic has grown in many countries (11,12). However, the frequency and characteristics of non-consensual sex remain to be elucidated, not only after but also before the COVID-19 pandemic in Japan to date, partly due to traditional social norms that discourage open discussions about sexual issues (13).

The purpose of our study was to report the incidence of non-consensual during the early stages of the COVID-19 pandemic in Japan. Additionally, we explored potential risk factors such as mental health conditions, economic changes, and concerns about the lockdown, and anxiety about the disease.

## METHODS

### Context

This cross-sectional study was conducted among a sample of approximately 2.2 million individuals from the general public who participated in a web-based self-reported questionnaire survey for the Japan COVID-19 and Society Internet Survey (JACSIS) project (14). The survey was administered by the well-established internet research agency, Rakuten Insight, Inc., which has been utilized in prior research studies (14,15). Data were collected from April 2020 to September 30, 2020. The prefectures where the selected individuals lived represented all prefectures in Japan, as indicated in previous studies (16).

### Patient and public involvement

No patients were involved in this study.

### Participants

The survey invitation was extended to 28,000 individuals out of 2.2 million registered with the survey agency, Rakuten Insight, Inc. Using a computer algorithm, study participants were selected through random sampling.

### Analysis Subjects

Respondents who self-identified as women in the questionnaire were included in our analysis. Among all the survey respondents totaling 28,000, the number of valid women participants for analysis was 12,809.

### Outcome Variable: Experience of Non-Consensual Sex

The focus of this study was to examine instances of non-consensual sex occurring between from April 2020 to 30 September 2020. Participants were not questioned about specific details but were instead asked to respond with a simple “yes” or “no” to indicate whether they had experienced such incidents.

### Exposure Variables Sociodemographic characteristics

As independent variables, demographics were included, encompassing gender, age groups (15–19, 20–29, 30–39, 40–49, 50-59, 60-69, 70-79 years), marital status (unmarried, married and widowed/separated), having children (none or more than one), educational attainment (high school educated, college educated or higher), employment status (any type of employment, unemployed), household income level, which was calculated as dividing the household income by the square root of household size (categorized by the tertials of household equivalent income (low, <JPY2.5 million/US$25 000/£16 667; intermediate, JPY2.5–JPY4.3 million/US$25 000–US$43 000/£16 667–£28 667; high, >JPY4.3 million/<US$43 000/<£28 667; unknown/declined to answer)), and smoking status (not smoker or current smoker).(14)

### Living Area Classification

Regarding living regions, all 47 prefectures were divided into the following three areas based on the date of the Declaration of the State of Emergency (DSE area) (17,18). “Designated as DSE on April 7, 2020”, “Designated as DSE on April 16, 2020”, and “Others”, “Specific alert designated on April 7, 2020” included Tokyo, Kanagawa, Chiba, Saitama, Osaka, Hyogo, and Fukuoka. “Specific alert designated on April 16, 2020” included Hokkaido, Ibaraki, Ishikawa, Gifu, Aichi, and Kyoto. “Others” comprised the remaining 34 prefectures. These categorization were determined by the Japanese Government using three indicators: the cumulative number of infected people, epidemiological trend, medical capacity and surveillance system (17). During the target period, people living in the relevant prefectures were advised to avoid unnecessarily outings, limit the use of entertainment facilities, and adhere to restrictions on economic activities (19).

### Fear of COVID-19 Scale

The Japanese version Fear of COVID-19 Scale (FCV-19S) was used to assess participants’ fear of the COVID-19 infection (20). This instrument comprises seven items rated on a 5-point scale (1 = “strongly disagree”; 5 = “strongly agree”). Consequently, the total score ranged from 7 to 35, with a higher score indicating a greater fear of COVID-19. Satisfactory internal consistency and validation of the scale were confirmed in the original seven-item scale (α= .82) (21).

### Personal Health and Economic Statuses

Based on previous research, an examination was conducted to determine whether health and economic statuses were associated with non-consensual sex (4). The health-related questions included the following: Self-rated health (good or other than good), “Any change in mental state in the last month compared to before January 2020 ?” (Worse, No change, Getting better, I do not know)”, Suicidal thoughts (Happened since before COVID-19 pandemic, First experienced during the COVID-19 pandemic or Never), Desire not to talk to anyone due to worries (Yes or No), Feeling isolated (Yes or No), Any cancellation of a family gathering due to the COVID-19 pandemic (Yes or No), Non-payment of salary (Happened since before COVID-19 pandemic, First happened during the COVID-19 pandemic, Never), and Lack of money to buy necessities of life (Happened since before COVID-19 pandemic, First Happened during the COVID-19 pandemic or Never).

### Statistical Procedure

To achieve the purpose of identifying the incidence of non-consensual sexual activity and its associated factors during the COVID-19 outbreak, two types of analysis methods were employed.

First, to determine the incidence rate, we divided the sample into two groups: those who responded “Yes” to the question of “Any non-consensual sex” and who did not. The number and percentage of individuals in these two categories, along with the χ-square test, were calculated for each independent variable. The number, percentage, and p-value of each item are described in Table 1.

**Table 1:** Demographics and Descriptive Statistics of Weighted Women Participants during the Five-Month from April to September 2020.

| Variable | Non-Consensual Sex April-Sept 2020 |  |
| --- | --- | --- |
|  | Total (n=12,809) |  |
|  | Yes<br>N (%) | No<br>N (%) |
| Total | 138 (1.1) | 12,671 (98.9) |
| Age |  |  |
| 15-19 | 8.0 (1.1) | 707 (98.9) |
| 20-29 | 37 (2.4) | 1543 (97.7) |
| 30-39 | 38 (2.0) | 1845 (98.0) |
| 40-49 | 25 (1.0) | 2395 (98.9) |
| 50-59 | 15 (0.7) | 2097 (99.3) |
| 60-69 | 15 (0.7) | 2127 (99.3) |
| 70-79 | 1.0 (0.1) | 1956 (99.9) |
| Marital status |  |  |
| Married | 88 (1.1) | 7929 (98.9) |
| Never married | 29 (0.9) | 3276 (99.1) |
| Widowed or separated | 20 (1.4) | 1467 (98.7) |
| Having children |  |  |
| No children | 56 (0.9) | 6006 (99.1) |
| At least one or more | 99 (1.5) | 6648 (98.5) |
| Educational attainment |  |  |
| High school or lower | 54 (1.0) | 4261 (99.0) |
| College/university or graduate school | 83 (1.2) | 7136 (98.9) |
| Employment status |  |  |
| Unemployed | 43 (0.7) | 6259 (99.3) |
| In employment | 95 (1.5) | 6412 (98.5) |
| Smoking status |  |  |
| Not smoker | 117 (1.0) | 11404(99.0) |
| Current smoker | 20 (1.6) | 1268 (98.4) |
| Household Income (million JPY) |  |  |
| Low | 53 (1.7) | 3144 (98.3) |
| Moderate | 43 (1.3) | 3226 (98.7) |
| High | 26 (1.0) | 2669 (99.0) |
| Unknown | 14 (0.4) | 3632 (99.6) |
| Region |  |  |
| Other | 86 (1.3) | 6628 (98.7) |
| DSE (April 16-) | 16 (0.8) | 2117 (99.2) |
| DSE (April 7-) | 31 (0.9) | 3568 (99.1) |
| Fear of COVID-19<sup>†</sup> |  |  |
| 7–15 | 41 (1.2) | 3402 (98.8) |
| 16–20 | 26 (0.6) | 4173 (99.4) |
| 21–25 | 34 (1.0) | 3405 (99.0) |
| 26–35 | 37 (2.1) | 1691 (97.9) |
| Self-rated health |  |  |
| Other than good | 94 (1.3) | 7349 (98.7) |
| Good | 44 (0.8) | 5322 (99.2) |
| Mental state change in the last month compared to before January 2020 |  |  |
| Worse | 59 (2.2) | 2662 (97.8) |
| None | 56 (0.6) | 8998 (99.4) |
| Better | 19 (3.3) | 547 (96.7) |
| Unknown | 4 (0.7) | 500 (99.3) |
| Suicidal thoughts |  |  |
| Yes, since before COVID-19 | 43 (4.0) | 1039 (95.1) |
| First happened | 20 (3.5) | 539 (96.5) |
| Never | 75 (0.7) | 11093(99.3) |
| Desire not to talk to anyone due to worries |  |  |
| Yes | 76 (4.0) | 2487 (97.0) |
| No | 62 (0.6) | 10184 (99.4) |
| Feeling isolated |  |  |
| Yes | 74 (3.7) | 1925 (96.3) |
| No | 64 (0.6) | 10747 (99.4) |
| Cancellation of a family gathering due to the pandemic |  |  |
| Yes | 10 (0.6) | 1730 (99.4) |
| No | 127 (1.2) | 10940(98.9) |
| Non-payment of salary |  |  |
| Yes, since before COVID-19 | 15 (10.3) | 133 (89.7) |
| First experience | 8 (10.4) | 67 (89.6) |
| Never | 115 (0.9) | 12471 (99.1) |
| Lack of money to buy necessities |  |  |
| Yes, since before COVID-19 | 35 (4.8) | 692 (95.2) |
| First experience | 22 (6.1) | 340 (94.0) |
| Never | 81 (0.7) | 11639 (99.3) |

<sup>†</sup> Classification of the Fear of COVID-19 scale scores refers to previous studies (22).
All estimates account for survey weights.

Second, a multivariate logistic regression analysis was conducted to identify potential predictive factors in the occurrence of non-consensual sex. This analysis encompassed all the aforementioned exposure variables, and odds ratios with 95% confidence intervals (CIs) were estimated (Table 2). As a sub-analysis, the incidence of suicidal ideation by age was also calculated.

**Table 2:**
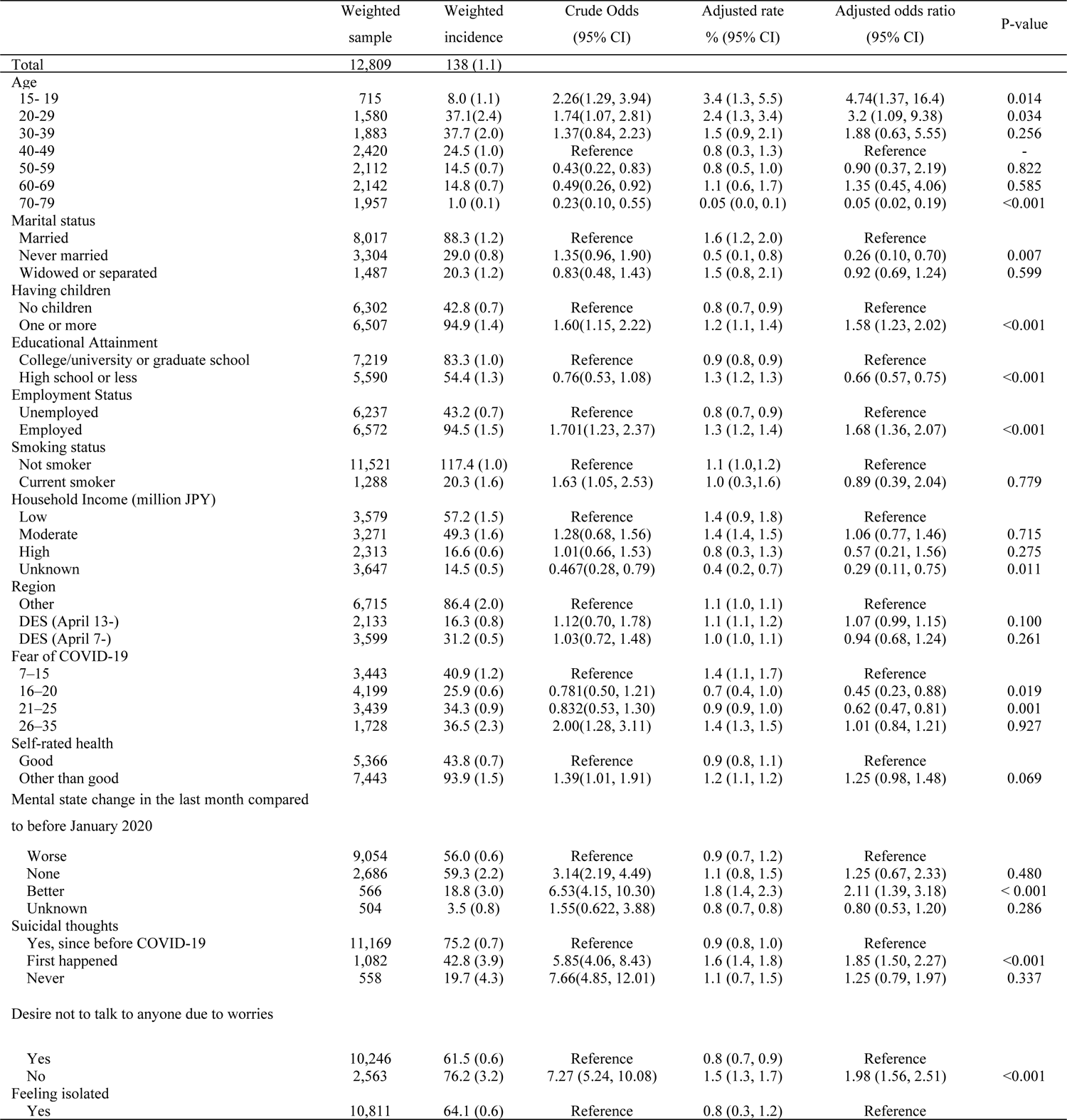
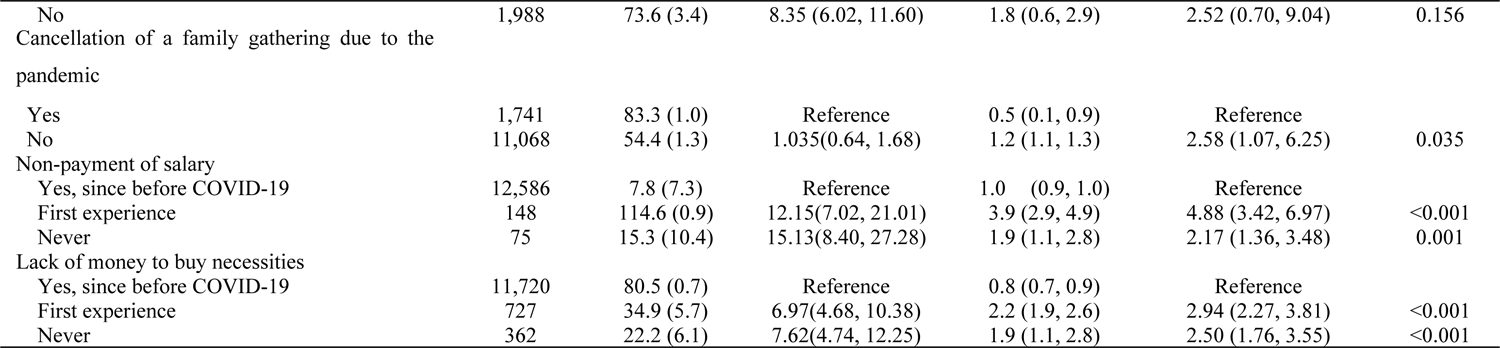
Factors Associated with Non-Consensual Sex Among Weighted Participants During the-Five-Month from April to September 2020.

It should be noted that because the characteristics of the participants in this Internet survey may differ from the general population (i.e., individuals with low digital skills might be excluded), a weighted analysis was performed to minimize differences in demographics, economic status, and health-related characteristics. This approach aimed to approximate national estimates from a nationally representative survey. Statistical significance was defined as p<0.05. The data were analyzed using STATA V.16.1 (Stata Corp, College Station, Texas, USA).

### Ethics

This study adhered to ethical standards in accordance with the 1975 Declaration of Helsinki, as revised in 2013. Prior to participating in the online survey, all participants provided informed consent through a web-based consent form. The Research Ethics Committee of the Osaka International Cancer Institute reviewed and approved the study (No. 20084).

## Results

The total number of respondents, including both men and women, was 28,000. Out of these, 2,518 participants who provided invalid responses (e.g., the same number is used for all survey items; incomplete responses) and 12,673 men were excluded. Consequently, the final analysis sample consisted of 12,809 women, whose ages ranged from 15 and 79 years old.

Table 1 shows the characteristics of the weighted proportion of non-consensual sex among all participants. The overall weighted incidence rate for non-consensual sex during the five-month period (April to September 2020) was 1.1 %. The incidence rate of non-consensual sex was higher among individuals aged 15-19 years (1.1%) and 20-29 years (2.4%), which exceeded the rates in other age groups. Women with jobs (1.5%) had a higher incidence rate than those without jobs (0.7%).

There was no significant difference in the incidence rate of non-consensual sex among those living in areas designated as DSE (0.9 % on April 7, 2020, 0.8 % on April 16, 2020, 1.3% others). A higher FCV-19S (2.1%, [26-35]) was associated with a higher incidence rate compared to other scores (1.2% [score from 7 to 15], 0.6% [score from 16 to 20], 1.0% [score from 21 to 25]).

The incidence rate of participants who reported feeling “worse” (2.2%) or “getting better” (4.4%) before the COVID-19 pandemic was higher than those who “No change” (0.6%) and “I do not know” (0.7%). Participants who had suicidal thoughts (4.0% since before COVID-19, 3.5% first happened) had a higher incidence rate than those who had never experienced them (0.7%). Participants who reported feeling isolated (3.7%) had a higher incidence rate than those who had never felt isolated (0.6%). Additionally, participants who had experienced non-payment of salary (10.3% since before COVID-19, 10.4% first experience) had a higher incidence rate than those who had never experienced it (0.9%). Similarly, participants who had experienced a lack of money to buy necessities (4.8% since before COVID-19, 6.1% first experience) had a higher incidence rate than those who had never experienced it (0.7%).

Table 2 shows the factors associated with non-consensual sex among participants. Participants aged 15 to 19 years and those aged 20 to 29 years had higher odds of experiencing non-consensual sex compared to participants in other aging groups. (Aged 15-19 years old: OR 4.74, 95%CI 1.37 – 16.4, Aged 20-29 years old: OR 3.20, 95%CI 1.09 – 9.38). Employed women participants were associated with higher odds of experiencing non-consensual sex than unemployed participants (OR 1.68, 95% CI 1.36-2.07).

Participants who did not receive salaries or compensations were more likely to experience non-consensual sex than those who did: (experienced since before COVID-19) OR 4.88, 95% CI 3.42-6.97; (first experience during COVID-19) OR 2.17, 95% CI 1.36-3.48. Similarly, participants who could not afford to buy life necessities were more likely to experiencing non-consensual sex than the others (OR 2.5, 95% CI 1.76-3.55).

Lastly, participants who had suicidal thoughts were more likely to experience non-consensual sex than those who did not: (since before COVID-19: OR 1.85, 95% CI 1.50-2.27). Participants with reported feeling isolated were more likely to experience non-consensual sex than those who did not (OR 1.98, 95% CI 1.56-2.51). The result of the sub-analysis showed suicidal ideation was present in 20% of all women aged between 15 and 19 years.

**Figure 1:**
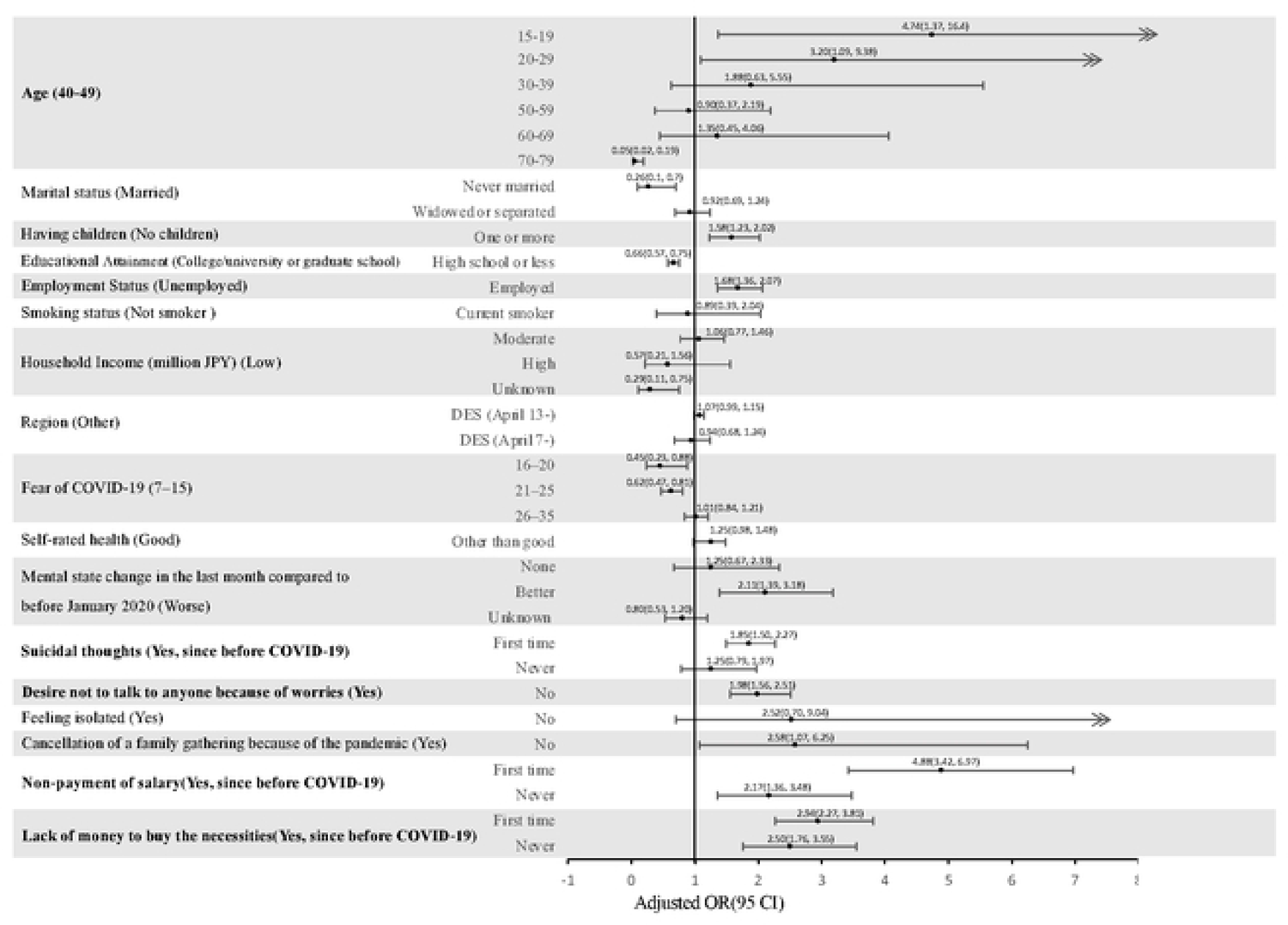
Forest plot illustrating incidence rate of non-consensual sex for each factor (reference) among the participants. Note: Significant positive factors associated with non-consensual sex included the age group "Aged 15-29 years old", as well as worsened mental or economic status were associated with an increased likelihood of experiencing non-consensual sex.

## Discussion

This study assessed the incidence of non-consensual sex in Japan during COVID-19 and explored associated factors while applying weighing to mitigate biases related to online participation. The overall rate of participants reporting non-consensual sex was 1.2% over a five-month period from April to September 2020. Factors associated with experiencing non-consensual sex among participants included ages between 15 and 29, employment status, deteriorating mental health condition, suicidal thoughts, a desire not to talk to anyone, feeling isolated, and challenging economic circumstances.

In Japan, research related to sexual matters is limited. We believe this limitation stems from the fact that sexual topics are often considered taboo with Japanese education institutions, and sexual education in Japan lags behind that in Western countries (22–24). Additionally, the understanding of obtaining sexual consent, which is essential in all sexual activities, lags behind compared to countries that are known to be advanced in sex education in some of Europe countries (13,22,25). For instance, the Netherlands had sex education compulsory from elementary school, and Denmark was one of the European countries that criminalized non-consensual sexual acts in 2020 at the time of our survey. Despite the efforts in Europe, conducting such a comprehensive study on a large scale was of significant importance as it ventured into uncharted territory in Japan.

Globally, concern arose about an increase in sexual violence against women around April 2020, during the early stages of the COVID-19 outbreak. However, specific incidence data were limited (12,26). While some countries reported an uptick in sexual violence cases (e.g., South Africa, Bangladesh (12,27)), others reported a significant decrease after implementing lockdown measures, as seen in Australia and Canada (28,29). In Japan, the available data included two key pieces of information from the police office: the reported prevalence of sexual violence among women, which stood at 6.9% (30), and a notable increase in the number of sexual assault victims seeking consultations, approximately 1.2 times higher in 2020 compared to 2019 (31,32). However, the incidence rate had not been thoroughly investigated during the COVID-19 pandemic, resulting in a scarcity of information regarding the incidence of sexual violence against women (33). It is important to note that our study did not have access to a comparative dataset from a different time point to evaluate percentage change in the incidence of sexual violence. Nevertheless, we were able to fundamental data concerning non-consensual sex for the first time in Japan.

Approximately 80% of the women who reported experiencing non-consensual sex fell within the age range of 15 and 49 years old. Notably, the incidence of reported non-consensual sex dramatically decreased among women aged 50 to 79 years. There was a significant increase in the risk of non-consensual sex within the age group 15 to 29 years. This pattern is consistent with reports from the United States and Japan, where individuals in their late teens to early thirties are identified as the age group at the highest risk of experiencing rape (30,34).

It is crucial to recognize that young women who have been victims of sexual assault are also more likely to experience lifetime suicide attempts and post-traumatic stress symptom. Specifically, research indicates that teenage sexual trauma is strongly correlate with suicide attempts (35–38). Additionally, our sub-analysis results revealed that suicidal ideation was present in 20% of all women aged between 15 and 19 years. Based on the aforementioned findings and considerations, we firmly believe that measures to protect the younger generation are imperative (39). As mentioned previously, Japan falls behind comprehensive sexual education for the younger generation. We contend that it is necessary to incorporate detailed explanations and the factors and circumstances that lead to unwanted sexual experiences involving the non-consensual sex into Japanese school curriculum.

Concretely, employed participants were more likely to be in contact with others than those who are unemployed. This time, we have not specifically investigated the locations where non-consensual sex has occurred. Therefore, it can be said that employed women may become victims of non-consensual sex, potentially by someone within their workplace or close social circle. Indeed, reports from both Japan and the United States have indicated that non-consensual sex is most frequently perpetrated by individuals known to the victims (30,40). Employed people have an extra circle of known individuals from their workplace compared to unemployed people.

Intriguingly, women who did not receive a salary and lacked the financial means to purchase necessities were at an elevated risk of experiencing non-consensual sex. While this appears to be the opposite of the above finding suggesting that being employed could be associated with a higher odds of experiencing non-consensual sex, this aligns with previous studies showing a connection between deteriorating economic conditions and an increased incidence of sexual assault that have established a connection between deteriorating economic conditions and an increased incidence of sexual assault (41,42), which is consistent with our findings. There are two possible explanations for this association: First, worsening personal economic circumstances may lead women to engage in occupations with a higher risk of non-consensual sex, such as working in the sex industry. Second, women in precarious financial situation often experience psychological pressure to secure and maintain employment (42). Under such pressure, they may become psychologically vulnerable to sexual coercion from their employers in exchange for job security (42). Future research should explore people in these circumstances in depth to better under-explored areas in Japan.

While 66% of the participants lived in areas under DSE, our study did not find a significant association between non-consensual sex and living in the DSE area. However, we did identify a significant correlation between experiencing a moderate level of fear related to the COVID-19 pandemic (scoring 16-25 points) and instances of non-consensual sex. This suggests that participants who stayed at home more frequently might have become more vulnerable to non-consensual sexual experiences by their partner. In fact, non-consensual sex was more frequently reported as being perpetrated by partner during the COVID-19 pandemic, and there was an observed increase in the number of domestic problems during this period (31).

Based on the considerations outlined above, there is a heightened risk of non-consensual sexual activity occurred during the COVID-19 pandemic, particularly among young, impoverished women. Consequently, it is of utmost importance to identify and provide economic and psychosocial support to these vulnerable individuals during times of social disruption. Furthermore, a notable deficiency in the current landscape of Japan is the absence of comprehensive education regarding the concept of sexual consent. Specifically, there is an urgent need for thorough instruction emphasizing the importance of men obtaining consent from women before engaging in any sexual activities (43). This education should be disseminated at both the local and national levels, as it may necessitate a societal shift in awareness at all levels.

In a noteworthy development, on June 16, 2023, Japan amended the legal term “Forced Sexual Intercourse Crime” to “Non-consensual Sexual Intercourse Crime”, explicitly stipulating that any sexual activity without consent is now considered a criminal offense (44). We anticipate that this alteration will provide a means of support to victims who have endured their suffering in silence and were unable to report their experiences.

## Limitations

This study has several limitations. First, as it is a web-based survey, participants in an internet-based study may not be representative of the general population. To address this potential bias, we performed statistical adjustments. However, it is important to note that this method may not fully account for the differences between participants in an internet survey and those in a nationwide representative survey, which poses challenges to the generalizability of our findings. To mitigate this limitation, harmonizing our data with a major national and representative cross-sectional study would enable us to pool data and potentially adjust for the factor of “being a respondent in an internet survey”, as has been done in other JACSIS studies (45).

The second limitation is related to the cross-sectional nature of survey. This design does not allow us to establish causal relationships between the presence or absence of non-consensual sex and factors such as economic status or suicidal ideation (46).

The third limitation concerns the definition of “Non-consensual sex”. We did not explicitly seek a specific definition of non-consensual sexual activity, which may be vary by generations or community cultures (e.g., workplace community, religious community). Sex education in Japan is known to be under-developed, therefore respondents may not necessarily share a relatively standardized understanding of this concept. We did inquire about employment status, but we did not specifically ask about non-consensual sexual activity in the workplace. However, we observed a significant correlation between being employed and experiencing non-consensual sexual activity. Also, we did not inquire the place of experienced non-consensual sex such as workplace. We observed a significant correlation between being employed and experiencing non-consensual sexual activity. However, it was not possible to ascertain whether non-consensual sex had occurred in the workplace.

The Forth, as mentioned in the section on “Analysis Subjects”, our questionnaire only asked whether respondents self-identified as male or female. Therefore, among those who self-identified as female, there is a possibility that transgender women or individuals who identify as bigender may also be included. This follows the convention in Japan, where many national surveys still only inquire about male or female gender identities.

## Conclusion

Early intervention to prevent individuals from becoming victims of sexual harm should be extended to economically vulnerable and young women, especially during times of societal upheaval such as the COVID-19 pandemic. Additionally, Japan should prioritize the implementation of comprehensive education on the concept of sexual consent.

## Data Availability

All relevant data are within the manuscript and its Supporting Information files.

## Acknowledgments

None

## Notes

### Competing Interest Statement

TT reports personal fees from Medical Network Systems, MNES Inc., and Bionics co. ltd., outside the submitted work. AO reports personal fees from Medical Network Systems, MNES Inc., outside the submitted work. All remaining authors have nothing to disclose.

